# Using Machine Learning to assess Covid-19 risks

**DOI:** 10.1101/2020.06.23.20137950

**Authors:** Srikanth Muthya, Renjith S Nair, Thanga Prabhu Arokiaswamy, Anusha Prakash

## Abstract

**IMPORTANCE:** Identifying potential Covid-19 patients in the general population is a huge challenge at the moment. Given the low availability of infected Covid-19 patients clinical data, it is challenging to understand and comprehend similar and complex patterns in these symptomatic patients. Laboratory testing for Covid19 antigen with <u>RT-PCR</u> | <u>(Reverse Transcriptase)</u> is not possible or economical for whole populations.

**OBJECTIVE:** To develop a Covid risk stratifier model that classifies people into different risk cohorts, based on their symptoms and validate the same.

**DESIGN:** Analysis of Covid cases across Wuhan and New York were done to identify the course of these cases prior to being symptomatic and being hospitalised for the infection. A dataset based on these statistics were generated and was then fed into an unsupervised learning algorithm to reveal patterns and identify similar groups of people in the population. Each of these cohorts were then classified and identified into three risk levels that were validated against the real world cases and studies.

**SETTING:** The study is based on general population.

**PARTICIPANTS:** The adult population were considered for the analysis, development and validation of the model

**RESULTS:** Of 1 million observations generated, 20% of them exhibited Covid symptoms and patterns, and 80% of them belonged to the asymptomatic and non-infected group of people. Upon clustering, three clinically obvious clusters were obtained, out of which the Cluster A had 20% of the symptomatic cases that were classified into one cohort, the other two cohorts, Cluster B had people with no symptoms but with high number of comorbidities and Cluster C had people with few leading indicators for the infection with few comorbidities. This was then validated against 300 participants whose data we collected as a part of a research study through our Covid-research tool and about 92% of them were classified correctly.

**CONCLUSION:** A model was developed and validated that classifies people into Covid risk categories based on their symptoms. This can be used to monitor and track cases that rapidly transition into being symptomatic which eventually get tested positive for the infection in order to initiate early medical interventions.

## INTRODUCTION

Covid19 has surprised the world with its infectivity and rapid spread globally causing massive loss of life and livelihoods. The right way to tackle this pandemic is to act quickly in identifying those at risk and treat patients early. Identifying and tracking symptoms of Covid infected patients is challenging today as new insights of its etiologic, pathology, public health impact, epidemiology, treatment options, vaccination etc. are emerging continuously with its global spread.

Machine learning has been extensively used in biomedical and medical sciences today to help in improving hospital outcomes, by effective early interventions that lead to improved prognosis. Data can be a powerful tool to analyse, interpret and build predictive models around them to support improved health care, if validated and analysed rightly. A scientific approach of using these techniques, can perfectly complement the clinical diagnostic and treatment protocols.

Getting access to datasets that capture the trends in the general population from being healthy to acquiring the infection and in-hospital prognosis phase is quite challenging and isn’t open source for the public due to obvious security and privacy concerns at the moment. Nevertheless, current investigations and studies are available that encapsulate most of the common statistics and symptoms of Covid patients. Using this, our proposed method captures these statistics along with some clinical background and generates a dataset on which we intend to apply an unsupervised learning algorithm to identify patterns and classify them into risk cohorts.

In predictive modelling, the term “Unsupervised Learning” refers to instances where the data does not have a label associated with it. Getting labels on data can be a very expensive process in terms of money, time and manpower. In such cases the knowledge is inferred from the data itself by applying clustering algorithms to find hidden, similar patterns and groups by some exploratory analysis. In our method, we have tried to infer patterns in different cohorts of people and label their Covid risk levels through analysis and further validation of the same.

In cases where the data isn’t available, one proven method in the healthcare space is to generate faux data through good clinical reasoning and validation. The data set is usually generated using a logic based algorithm that captures human knowledge about the subject along with current research and studies with some evidence. Covid based research has evidently increased since the pandemic has struck and related resources are available extensively today, and this method has tried to capture these studies into an interpretable form for analysis and categorization of different risk cohorts that were validated against current data. This model can be used to identify risk levels, based on which cohort they belong to or transition into, over a period of time.

## RELATED WORK

Creating synthetic datasets in healthcare is predominantly increasing because of the existing challenges in healthcare systems to record information in EHR and EMR formats and even if this isn’t a hindrance, security and privacy controls laws on these data are very stringent that it becomes hard to get access to. Nevertheless, synthetic data sets can be evolved to a better real world representation without compromising on the quality of the clinical information but also can help avoiding privacy clauses and concerns around them. ^[1]^

One such notable example is GAN’s (Generative Adversarial Network) in the deep learning research space that generates completely synthetic data with real world logics. medGAN(Choi et al)^[2]^ is an algorithm that generated realistic synthetic EHR’s that were high dimensional and discrete in nature using GAN’s and autoencoders. The RCGAN^[3]^is another interesting work that generated high dimensional realistic synthetic time series datasets using Recurrent GAN’s. Most of the GAN techniques applied in healthcare settings had some or very little real world data that was fed into GAN’s which isn’t the case with our problem statement. The availability of COVID patient records at absolutely zero today. Two drawbacks of using GAN’s are validation and poor interpretability in assessing why some samples are created and this makes it hard to implement.

Laura et al.(2018)^[4]^ used Naive Bayes clustering methods to generate realistic datasets taking MIMIC III as a baseline and had much better results compared to medGAN^[2]^.

One very similar approach as ours as is, of Chen et al.(2019)^[5]^which generated more than a million “synthetic residents” by an algorithm named Synthea, also called as Synthetic mass that represented residential population around Massachusetts, USA and mimicked the statistics of the population including their demographics, vaccinations, medical visits and comorbidities. This was also compared and validated with the original population around the city.

Another notable work is of Harvard Dataverse, which has 10,000 completely synthetic datasets of patients generated from software called Synthea that was mentioned previously.^[7]^

Mahmoud et al.(2017) used K-means clustering to predict patient outcomes in elderly patients^[19]^.Hany et al.(2019) have summarized how clustering techniques and pattern identification in AD patients(Alzheimer’s disease), from early to last stages of the disease can be effective in healthcare^[20]^. Lio (et al.) applied clustering techniques to find patterns in end stage renal disease patients who initiated Hemodialysis ^[22]^.

## PROPOSED METHOD

### I. PROBLEM ASSESSMENT

Coronavirus is known to progress in some infected patients, affecting the vital organs of the body rapidly. Not everybody will experience similar symptoms, it varies from person to person and a majority remain asymptomatic. It becomes challenging to identify such patterns in the general population. If the population is tracked for symptoms constantly, monitored and assessed for infection, then identifying people who are likely to be infected can initiate early interventions.

### II. FAUX DB GENERATION

To understand complex patterns and symptoms in infected cases, A real world dataset explaining clinical conditions during the asymptomatic case is required to do any research and build predictive models around it. Obtaining such historical clinical information of cases can be very expensive, time consuming and sensitive to be made open source to the public. Often, even if this kind of data existed, clinical records have missing gaps and acute information that aren’t adequate enough to draw conclusions from. With this challenge of obtaining clinical data by conventional methods, generating a “synthetic but convincing” dataset to understand the patterns in symptomatic cases with current evidence and studies is the need of the hour. Studies have shown that generating synthetic clinical dataset is a promising and a plausible approach to take in such scenarios that can solve current problems.

Using statistical studies related to Covid infected cases across cities of Wuhan, China and New York, USA a dataset was generated that fit across these populations and describes them well enough to work on. Although the statistics of infected cases across the globe are contrasting to one another, an effort was made to capture the recurring patterns and similarities in both the cities that normalises this difference to an acceptable level. The reason why cases across these cities were chosen in particular, is primarily due to the fact that they have the most number of cases with a huge population and validation of our dataset would use this as a baseline and for future studies.

Covid19 In-hospital admission information was considered from the period March 1, 2020 until April 4, 2020 from an investigation conducted in New York^[8]^, which was the epicenter of Covid cases in the United States. This investigation consisted of a total of 5700 participants who were diagnosed with the infection and had received treatment for the same. Statistics of these patients included the comorbidities, symptoms, age, gender, race and more. Similarly, the characteristics from Wuhan, which happens to be the world’s epicentre for the virus, were studied, from the period December 8,2019 to March 8,2020.^[9]^ This study investigated the trends in the spread of the virus, and symptoms. They were studied across different cohorts of population that were classified into mild, moderate and severe with respect to the infection. It also captured similar characteristics of that of the former study mentioned.

Both of these studies were compared against each other in terms of infected population’s statistics and were found to be contrasting at few places with different numbers in demographics like gender and age. The common characteristics were found to be comorbidities and the early symptoms of the infection.

We tried minimising these differences and came up with numbers that equated instances from both the investigations and fairly generalised the infection trends and symptoms for a general population. We do not intend to build a universal data set that represents the global population. Our interest is to capture major symptomatic and infection prone populations based on the studies till date and simulate the same.

The idea behind generating synthetic dataset begins with exploiting freely available information regarding the statistics, prevalence and incidence of this infection. From these statistics, we can get a fair picture regarding the demographics, and prevalence of symptoms and comorbidities in the infected population. The synthetic data was generated by GRiSER’s method^[21]^.Fig 1 explains the approach to build our dataset from open source information and clinical knowledge.

**Fig 1.**
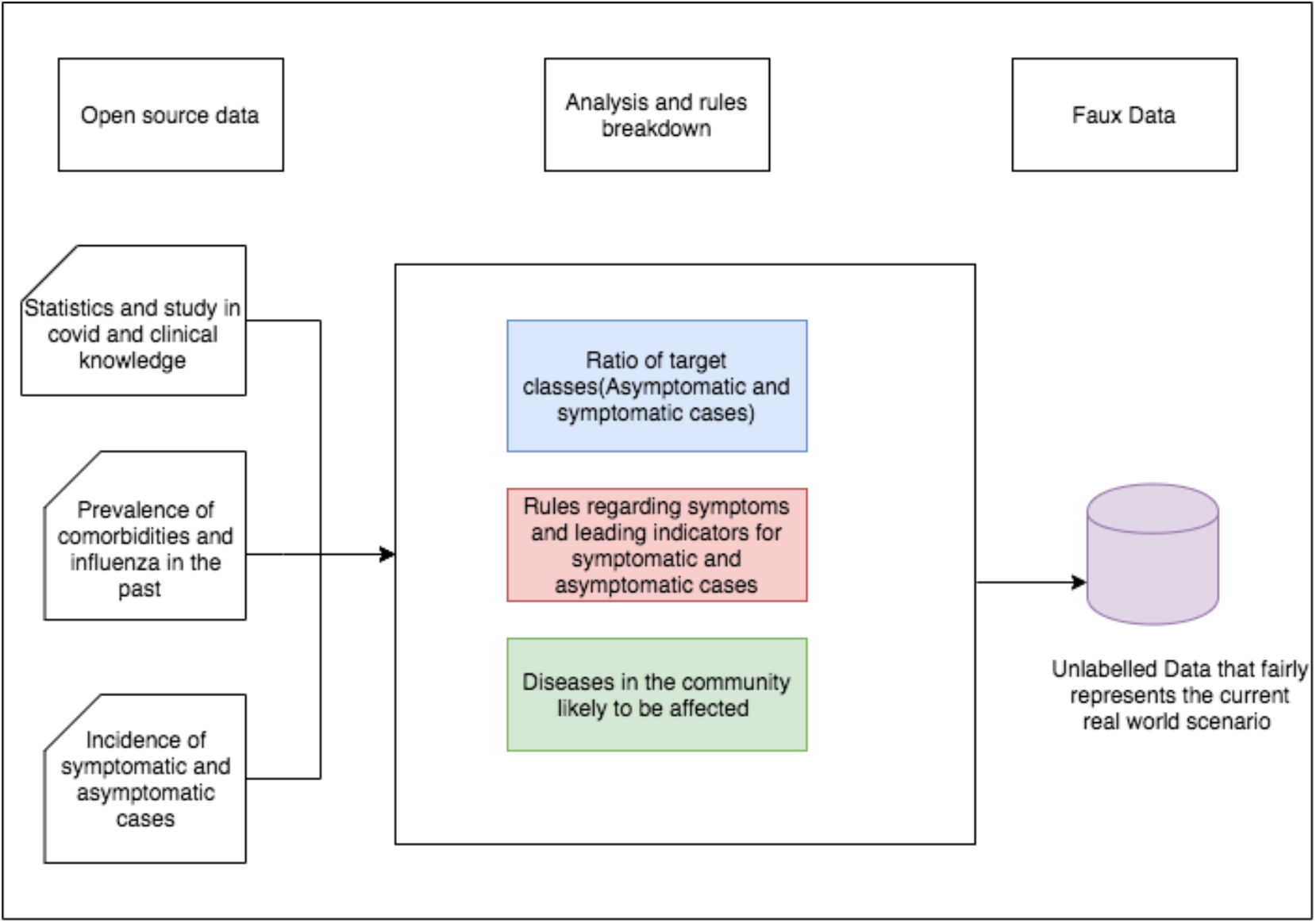
Approach to generate the faux dataset using available data and studies.

**Fig 2.**
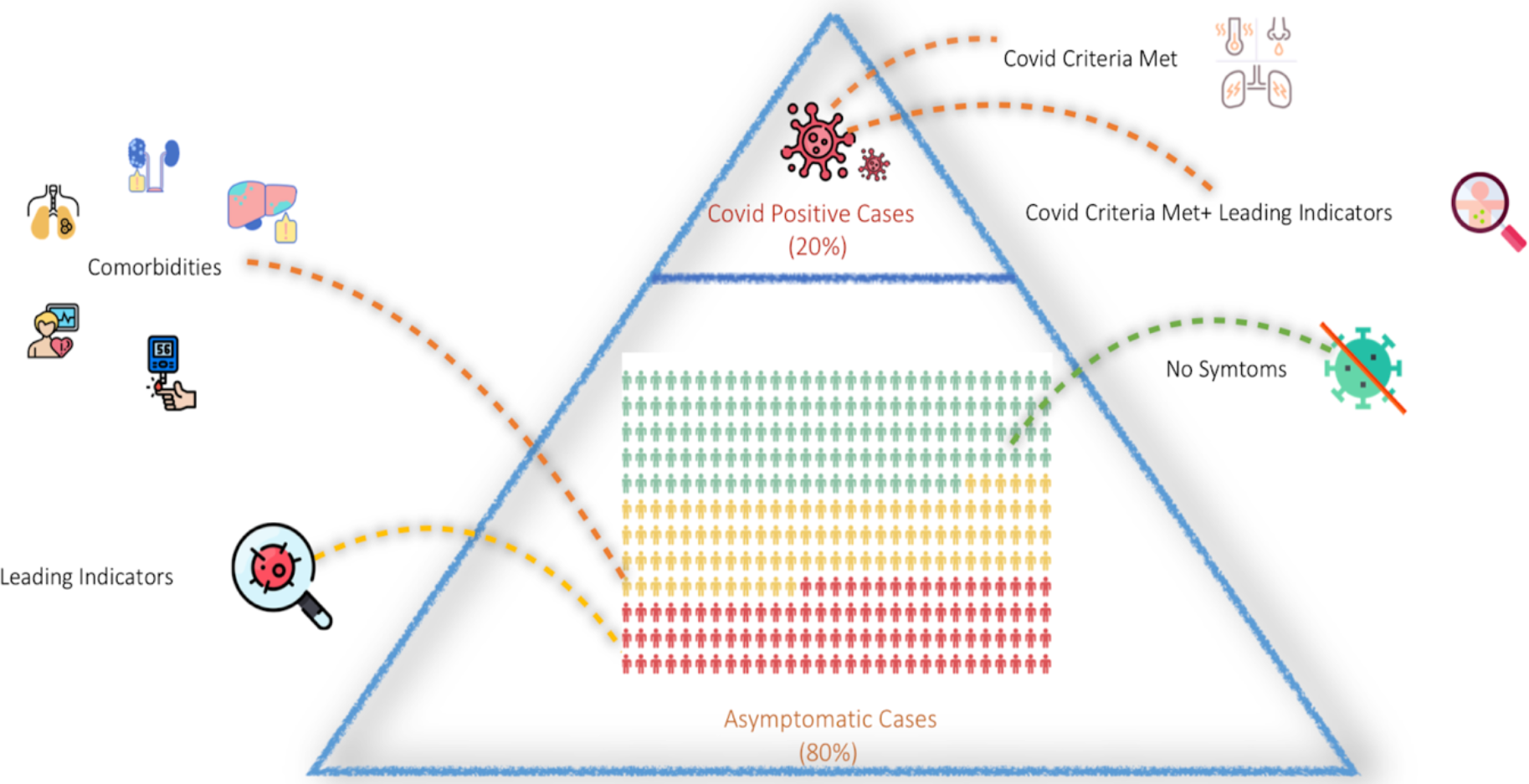
Representation of the real world Covid cases, prevalence disease statistics of Covid symptoms and leading indicators for the infection.

**Fig 3.**
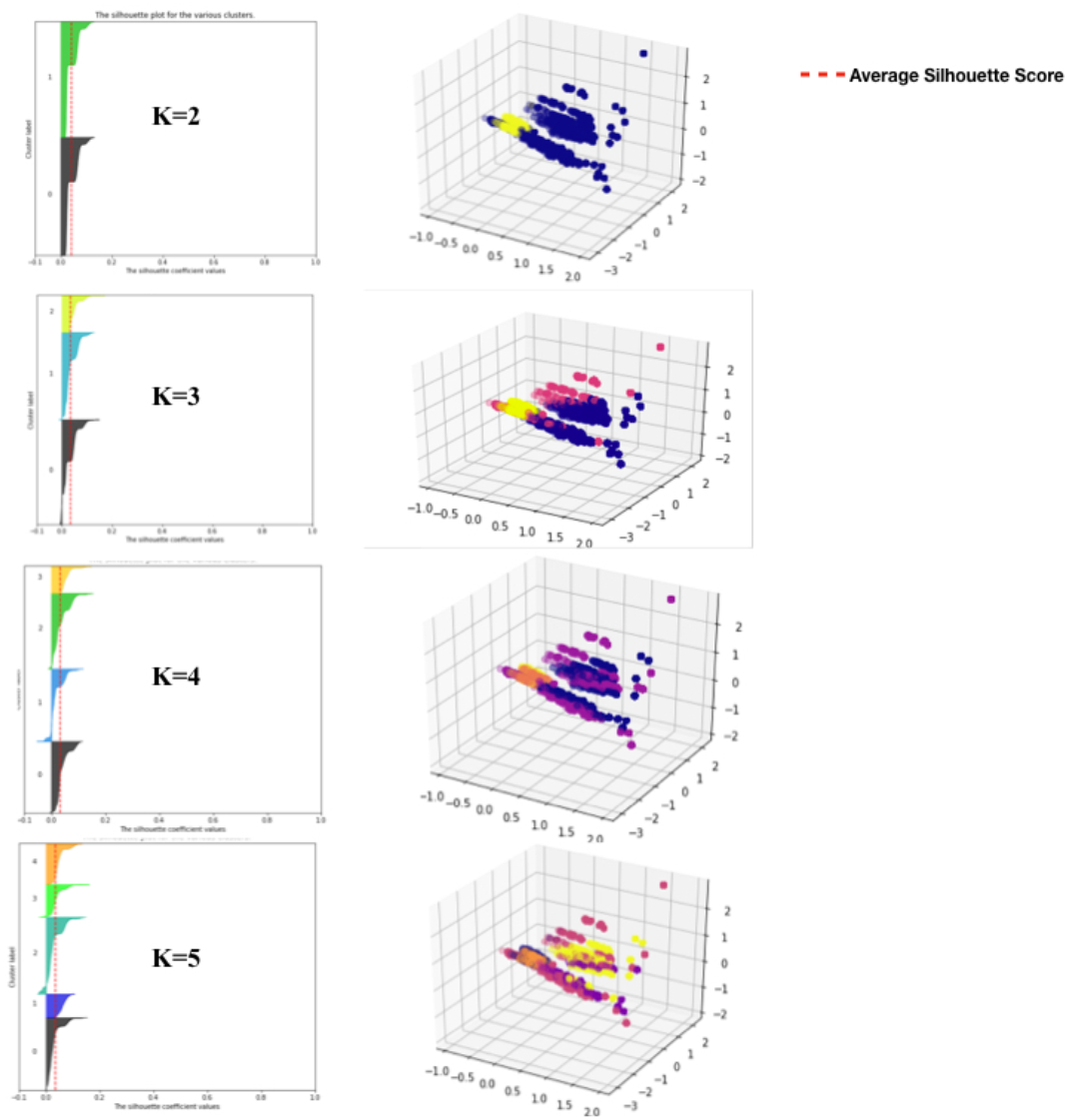
Silhouette plots and clusters formed on different iterations and values of K. The clusters are visualised post MCA(Multiple Correspondence Analysis) and were projected on a 3-Dimensional space. K=3 Seems to the reasonable choice given the cost and silhouette scores.

### III. DATA DESCRIPTION

The features we considered were symptoms observed in infected patients and comorbidities. We did not consider age as a feature since we believe “covid19 is a de novo disease. initially thought to affect elders predominantly. With time, other age groups were also affected but mechanism is poorly understood. Age does not appear to be a primary factor in getting infected or disease progression”. We define “Covid criteria met” definition based on higher incidence and prevalence of certain symptoms associated with this virus that is likely to be experienced by the host during the initial stages of the infection.[9] We also identified few leading indicators or signs that were likely to occur in some symptomatic populations. For example, Diarrhea, nausea, conjunctivitis and loss of taste and smell[11] were found to be in the very early stages of the infection. Also, Travel history along with flu-like symptoms can be a strong indicator of the infection. The Covid criteria met definition was curated from observed symptoms in infected patients globally and coronavirus studies.^[8][9][10]^ Table 1 explains the Covid criteria met definition and Table 2 explains the features we have identified and considered as a part of our study.

**Table 1.** Features and rules considered for Covid criteria met condition.

| Covid criteria met definition |
| --- |
| Presence of Fever<br>AND<br>Dry Cough<br>AND<br>Shortness of breath<br>AND<br>Fatigue<br>AND<br>Any 1 of the comorbidities<br>( Heart disease, Diabetes, Lung disease, Hypertension, Kidney ) |

**Table 2.** Features identified as Leading Signs for the infection.

| Leading Indicators for Covid |  |
| --- | --- |
| Chills | Loss of Taste and Smell |
| Muscle Ache | Chest Pain |
| Sore throat | Nausea |
| Conjunctivitis | Headache |
| Vomiting | Swollen Lymph Node |
| Diarrhea | Wheezing |
| Travel history | Low Immunity |
| Runny nose |  |

We generated over 1 Million records that captured the above statistics and demographics with categorical information(Boolean values). The description of the data generated is explained in Table 3.

**Table 3.** Data Description with all the features.

| Data Characteristics | n (%) |
| --- | --- |
| Total Number | N=10,00,000 |
| **Gender** |  |
| Female | 4,87,000 (48.7%) |
| Male | 5,13,000 (51.3%) |
| **Symptoms** |  |
| Fever | 232047 (23%) |
| Dry Cough | 231724 (23.1%) |
| Shortness of Breath | 232392 (23.2%) |
| Fatigue | 240290 (24.0%) |
| Chills | 180308 (18.0%) |
| Muscle Ache | 201907 (20 %) |
| Sore throat | 201690 (20.1%) |
| Conjunctivitis | 96197 (9.1%) |
| Vomiting | 164063 (16%) |
| Diarrhea | 164236 (16.4%) |
| Travel history | 163984 (16.3 %) |
| Runny nose | 179910 (17.9 %) |
| Loss of Taste and Smell | 125729 (12.5 %) |
| Chest Pain | 107788 (10.7 %) |
| Nausea | 199479 (19 %) |
| Headache | 179889 (17.9 %) |
| Swollen Lymph Node | 136131 (13.6 %) |
| Wheezing | 210417 (21%) |
| Low Immunity | 519633 (51.9 %) |

**Table 3. Data Description with all the features.**
| Comorbidities |  |
| --- | --- |
| Heart Disease | 246000 (24.6%) |
| Lung Disease | 331400 (33.1%) |
| Kidney Disease | 117243 (11.7 %) |
| Hypertension | 506743 (50.6 %) |
| Diabetes | 343600 (34.3 %) |

### IV. CLUSTERING APPROACH

Clustering analysis is used on unlabelled dataset to learn different cohorts and patterns in the data. Most popular clustering algorithm used in healthcare applications is the K-means clustering algorithm. This is used when we have numerical and continuous clinical data^[12]^. K-means algorithm^[13]^ groups similar data points together and identifies the underlying patterns within them. It uses distance based metrics to group data into K different clusters by calculating K different centroids(An imaginary location that represents the center of the cluster) and assigning every data point to the nearest centroid.

Applying K-means on our data does not make sense since, Euclidean distance isn’t the right distance metric when we have a dataset with categorical features. Rather, we need to capture the dissimilarity measure between our data points. Here is when we use the K-Modes algorithm, which is a slight extension of K-means, except that it quantifies the dissimilarity between two data points rather than compute distances.^[14][15]^ The K-Modes algorithm can be explained on a high level

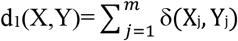

where (X_j_,Y_j_) is defined as

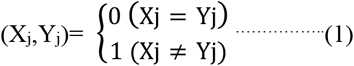

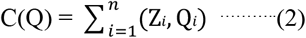

where Z is the categorical variables ranging from A_0_,A_1_…A_n_ and Zi is the i^th^ element and Qi is near cluster center.

**Listing 1**. Explains high level algorithm for K-modes Clustering.

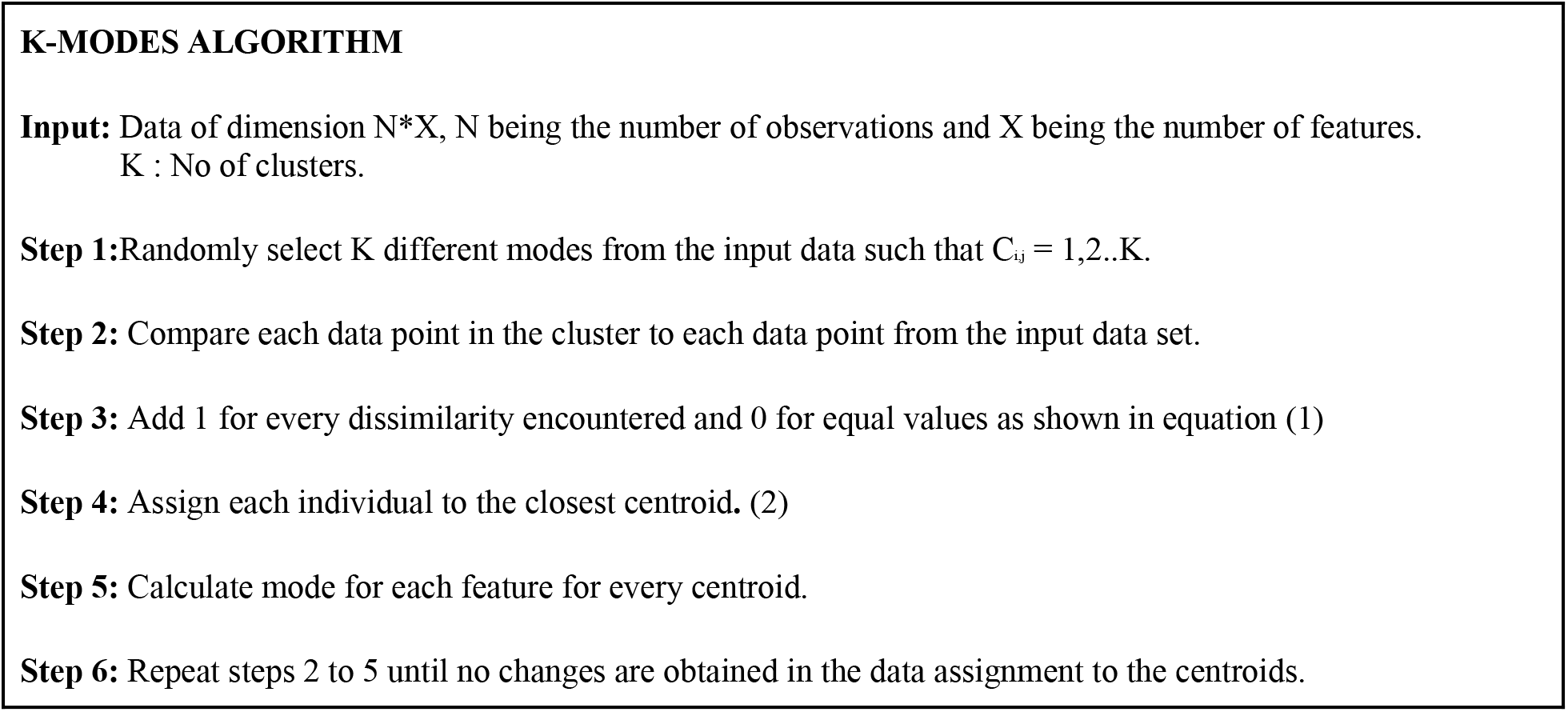

#### IV a. CHOOSING OPTIMAL K VALUE

Choosing the optimal K or number of cluster values is a very important step in clustering. We applied silhouette scoring and cost analysis to arrive at our K value. Silhouette score is a measure of similarity between a data point and its own clusters^[16]^. The best value is supposedly close to 1 and negative values would indicate data points assigned to wrong clusters. The K is chosen based on the high silhouette coefficient value obtained after iterating through various K values. The silhouette coefficient is usually measured based on the Euclidean distance. Given that our data has a non-gaussian and discrete distribution, we apply hamming distance instead of the Euclidean distance.

Post the silhouette score and cost analysis on various K values (Refer Table 4). We found K=3 would be an optimal number for our objective. Visualisation of clusters is still a question of research today, Nevertheless, to get a fair idea and validity of our algorithm, we applied MCA(Multiple Correspondence Analysis) on the data and applied it in order to project it on a 3D space. Multiple Correspondence Analysis tries to identify associations in multiple categorical variables.^[18]^ The parameters Init was set to Huang, K was set to 3 and n_init was set to 6 in the K-modes parameter selection process.

**Table 4.** Cost values on different K-values.

| K Value | Cost |
| --- | --- |
| 2 | 4442956 |
| 3 | 3946176 |
| 4 | 4176818 |
| 5 | 3007670 |

**Table 5**. Explains the statistics obtained on each cluster. Next step was to identify the risk groups among these cohorts. Cohort risk identification was made based on clinical knowledge and evidence of Covid studies. From the inferences we drew from the above clusters we assigned subjective risks namely Low, Medium and High to Cluster B, Cluster A, Cluster C respectively.

Cluster C happens to contain the symptomatic group of people with same exhibiting characteristics of the ones that were investigated and observed globally till date. Hence, it is identified as a High risk group that is likely to be symptomatic for Covid.

**Table 5.** Cluster statistics and description after applying K-modes.

| Features | Cluster A prevalence n (%) | Cluster B prevalence n (%) | Cluster C prevalence n(%) |
| --- | --- | --- | --- |
| **N=10,00,000** | **6,01,458 (60%)** | **1,99,998 (19.9%)** | **1,98,544 (19.85%)** |
| **SYMPTOMS** |  |  |  |
| Fever | 25403 (4.2%) | 8102 (4.5%) | 198542(98%) |
| Chest Pain | 6603 (1.1%) | 2037(1.02%) | 99148(48%) |
| Chills | 31166 (5.18) | 9988(4.9%) | 59563(30%) |
| Headache | 60762(10.1%) | 19832(9.92%) | 99295(50%) |
| Nausea | 60726(10.1%) | 20097(10%) | 118656(59.8) |
| Diarrhea | 19036(3.16%) | 5921(2.96%) | 60601(31.1%) |
| Fatigue | 31713(5.27%) | 10038(5.02%) | 198539(98%) |
| Change in Taste or smell | 12758(2.12%) | 3929(1.96%) | 109042(54.9%) |
| Swollen Lymph Node | 12856(2.14%) | 3917(1.96%) | 79417(40%) |
| Shortness of breath | 25760(4.28%) | 8095(4.05%) | 198537(98%) |
| Cough | 25287(4.2%) | 7899(3.95%) | 198538(97.8%) |
| Wheezing | 61138(10.2%) | 20000(10%) | 129279(65%) |
| Sore throat | 24000(3.99%) | 7942(3.97%) | 169748(85.5%) |
| Vomiting | 18893(3.14%) | 6067(3.03%) | 60612(32%) |
| Muscle Ache | 24030(4%) | 7986(3.99%) | 169891(85%) |
| Runny Nose | 30937(5.14%) | 9919(4.96%) | 139054(70%) |
| Conjunctivitis | 12720(2.11%) | 3950(1.98%) | 39727(21%) |
| Travel History | 19092(3.17%) | 6061(3.03%) | 138831(70%) |
| **COMORBIDITIES** |  |  |  |
| Diabetes | 207771(34.5%) | 68711(34.4%) | 67118(33.8%) |
| Heart Disease | 73861(12.3%) | 24071(12%) | 148068(74.6%) |
| Hypertension | 401465(66.7%) | 0(0%) | 105278(53%) |
| Kidney Disease | 13930(2.32%) | 4369(2.18%) | 34369(17.1%) |
| Lung Disease | 222371(37%) | 74660(37.3%) | 98944(49.8%) |
| Low Immunity | 401466(66.7%) | 0(0%) | 118167(60%) |

### V. INTERNAL VALIDATION

Internal validation of this model is essential to gauge the accuracy of it and test the sensitivity of the algorithm’s ability to profile a new data point into its right risk group. To perform this validation, we analysed data from an open source database that had high level summaries of the corona positive patients at the time of detection^[17][23]^. We used this information to simulate timelines for various scenarios from the onset of the symptoms till confirmed date. We simulated about 1500 observations for 50 patients based on the age group, symptoms developed over a timeline of 7 to 14 days. We essentially captured 3 major use cases that included, Covid symptoms with leading indicators, Covid symptoms without leading indicators, Flu like symptoms with travel history, all of these developed over 7 and 14 days. The objective was to identify if the model could distinguish between normal flu and Covid symptoms that had lower incidence in the general population.

Fig 4. Shows results of the validation when run through the covid risk model.

**Fig 4.**
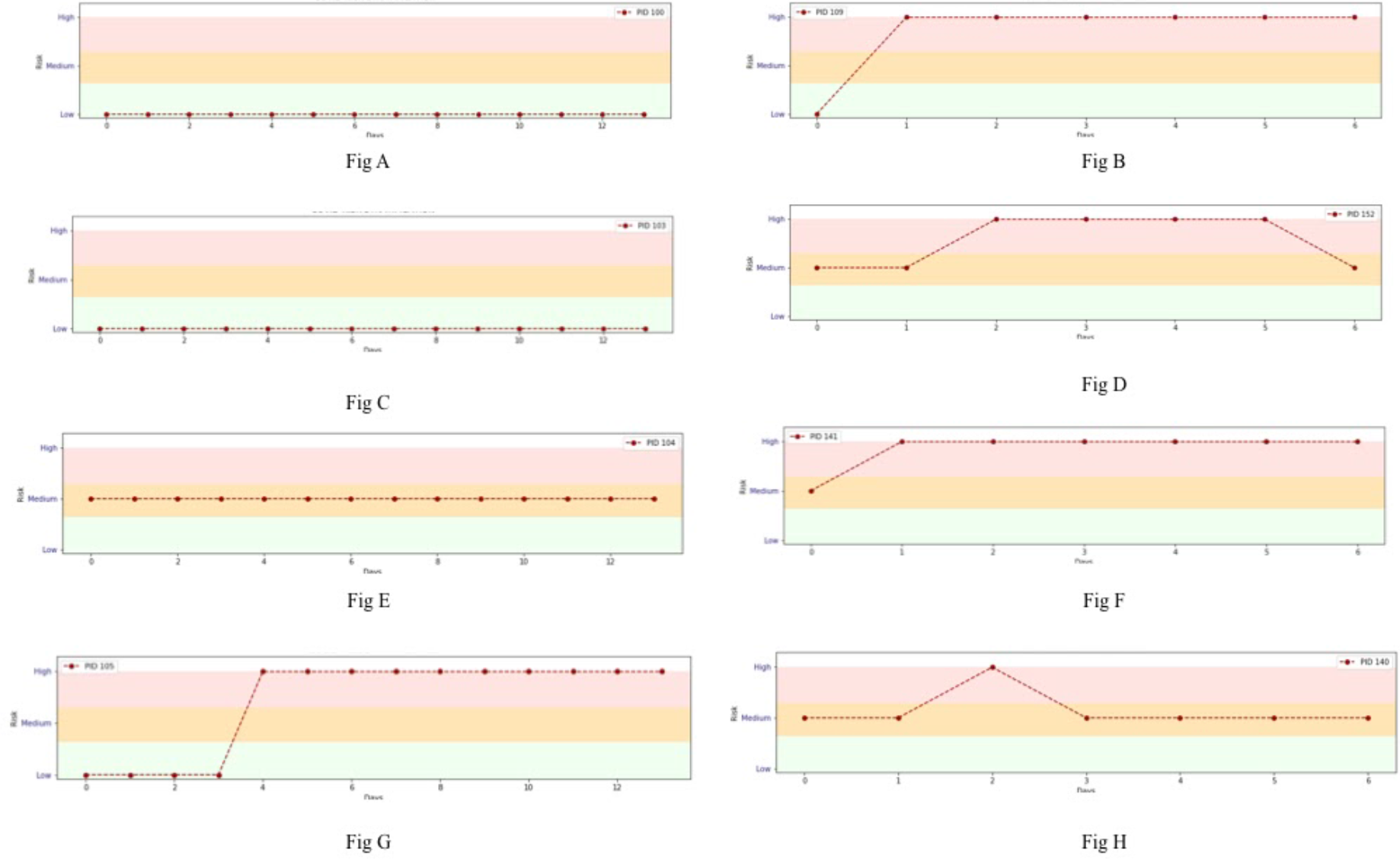
Graphs showing the risk trend for six different timelines from the subset of the validation set. Fig A is of a young, healthy person with no symptoms. Fig B is of a young person with covid criteria symptoms with Leading Indicators developed over a week. Fig C is of a young person with flu like symptoms(Not Covid criteria). Fig D is of an Old aged, Unhealthy(Comorbidities) patient with Hypertension, Diabetes, Covid criteria along with leading indicators developed over a week. Fig E is of a Young, Healthy person with travel history and low Immunity. Fig F is of a Middle aged, Unhealthy(Comorbidities) person with Lung Disease, Covid criteria with leading indicators developed over 1 week. Fig G is a Young, Healthy person with Covid criteria developed over 14 days. Fig H is a Middle aged, Unhealthy(Comorbidities), Hypertension, Heart disease, Covid criteria without leading indicators developed over 1 week.

The model was sensitive towards the Covid met indicator conditions and leading indicators, but there were few false positives in example cases like, the risk was “HIGH” when it gave a more weightage to conditions like low immunity and travel history. This is expected to improve when the model is re-trained on a bigger real world dataset that has complex correlations and patterns.

### VI. EXTERNAL VALIDATION

Covid19 Research Tool (C19RT) is a web application developed by the team at Cohere. The idea behind this web application is to collect data from individuals for our research study and track their symptoms to identify the risk of infection. This web application is for the public, and allows anybody to register and enter their symptoms at least once a day for a period of 30 days. With prior research on Covid symptoms from various sources and our clinical advisory board, we curated a questionnaire that was user friendly and targeted all levels of population. We released this application in the month of May and have collected (still collecting) about 1000 plus data points from 300 users. We have a privacy and security protocol in place to handle this collected information that is anonymised and run through the Covid model in the backend.

#### VI a. C19RT WORKFLOW

A new user registers into our application, after acknowledging the consent form. The designated user enters his symptoms at least once a day on the web application(can be accessed on any digital device). This information is then anonymised, run through the Covid risk algorithm which profiles this input with a similar group of people and gives out the group/ risk category the person belongs to. When a change in the risk trend is observed, an alert is emailed to the user. The entire application workflow is explained in Fig 5.

**Fig 5.**
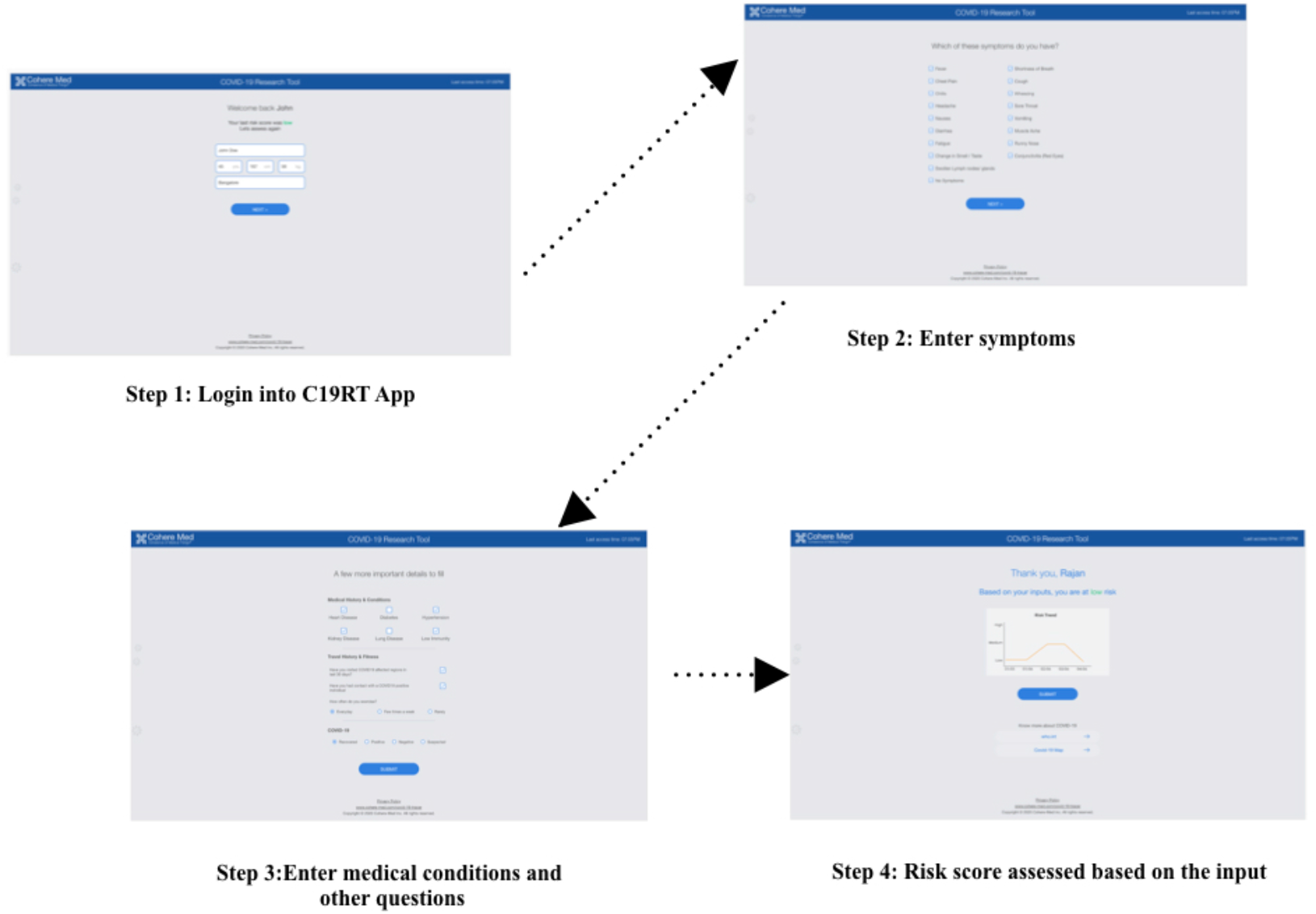
The C19RT workflow step-wise.

The collected data along with the risks were validated clinically and was compared against the targets generated for each of them manually with clinical knowledge. The classification report and confusion matrix of this validation is shown in Table.6 and fig 6 respectively.

**Table 6.** Risk Stratification analysis on the real world data.

| Clusters | Recall | Precision |
| --- | --- | --- |
| Cluster A (Medium Risk) | 72% | 82% |
| Cluster B (Low Risk) | 85% | 85% |
| Cluster C (High Risk) | 92% | 84% |

**Fig 6.**
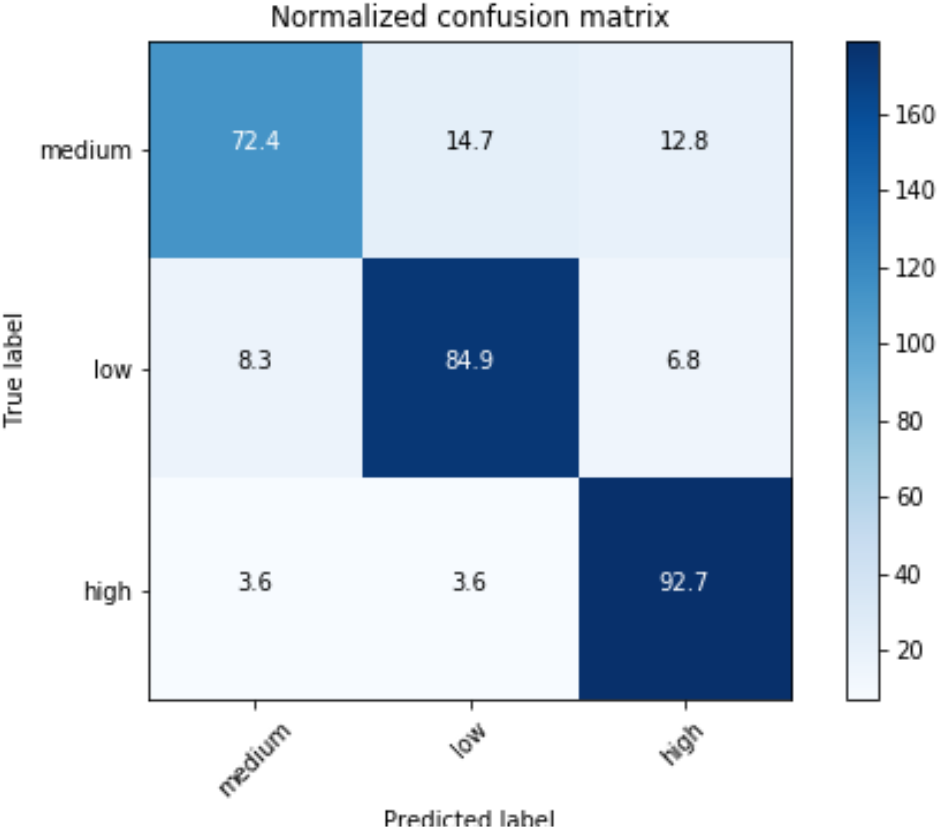
Confusion Matrix obtained on validation set.

## CONCLUSION

We were successfully able to validate and deploy this model. Our research tool at present uses this model to display a Covid risk score based on the user’s input.

Understanding Covid patterns in the symptomatic cases is still an ongoing challenge and subject of research. Although, there are few sets of rules or pointers that could indicate the presence of the infection, just a rule based solution wouldn’t be the right approach. Rule based decision systems tend to generate a lot of false positives and have a relatively low precision. While, significantly larger real world data is definitely the key to better insights, and results, our objective is to mimic the real world scenarios and identify patterns in order to catch these symptoms earlier. The earlier the treatment, better the prognosis. Unsupervised learning like clustering can be a powerful analysis in healthcare, because often in practice, clinicians tend to profile similar cases and conditions along with other dimensions of a patient to land at an informed decision. Clustering that way, is mimicking this concept, with only a superior ability to identify patterns across a huge dataset in terms of dimensions and size.

For the model to capture more complex correlation across the features and cohort patterns, our goal is to continue to collect relevant data from a larger population to improve the algorithm, learn better patterns and reveal insights that can help identify those with high risk of being infected with Covid19. This is just one part of the problem that we try to solve. A bigger challenge is identifying the asymptomatic cases that go unidentified and unnoticed, spreading widely in the population, described as community transmission by epidemiologists.

## Data Availability

This work used synthetic data generated from studies. For internal and external validation, open data sources were used.

https://www.kaggle.com/rupsikaushik/covid19-patientlevel-data

https://github.com/beoutbreakprepared/nCoV2019.

